# AI based pre-screening of large bowel cancer via weakly supervised learning of colorectal biopsy histology images

**DOI:** 10.1101/2022.02.28.22271565

**Authors:** Mohsin Bilal, Yee Wah Tsang, Mahmoud Ali, Simon Graham, Emily Hero, Noorul Wahab, Katherine Dodd, Harvir Sahota, Wenqi Lu, Mostafa Jahanifar, Andrew Robinson, Ayesha Azam, Ksenija Benes, Mohammed Nimir, Abhir Bhalerao, Hesham Eldaly, Shan E Ahmed Raza, Kishore Gopalakrishnan, Fayyaz Minhas, David Snead, Nasir Rajpoot

**Author notes:** Joint first authors (Mohsin Bilal, Yee Wah Tsang). Joint last authors (David Snead, Nasir Rajpoot).

## Abstract

Histopathological examination is a pivotal step in the diagnosis and treatment planning of many major diseases. To facilitate the diagnostic decision-making and reduce the workload of pathologists, we present an AI-based pre-screening tool capable of identifying normal and neoplastic colon biopsies. To learn the differential histological patterns from whole slides images (WSIs) stained with hematoxylin and eosin (H&E), our proposed weakly supervised deep learning method requires only slide-level labels and no detailed cell or region-level annotations. The proposed method was developed and validated on an internal cohort of biopsy slides (n=4 292) from two hospitals labeled with corresponding diagnostic categories assigned by pathologists after reviewing case reports. Performance of the proposed colon cancer pre-screening tool was evaluated in a cross-validation setting using the internal cohort (n=4 292) and also by an external validation on The Cancer Genome Atlas (TCGA) cohort (n=731). With overall cross-validated classification accuracy (AUROC = 0.9895) and external validation accuracy (AUROC = 0.9746), the proposed tool promises high accuracy to assist with the pre-screening of colorectal biopsies in clinical practice. Analysis of saliency maps confirms the representation of disease heterogeneity in model predictions and their association with relevant pathological features. The proposed AI tool correctly reported some slides as neoplastic while clinical reports suggested they were normal. Additionally, we analyzed genetic mutations and gene enrichment analysis of AI-generated neoplastic scores to gain further insight into the model predictions and explore the association between neoplastic histology and genetic heterogeneity through representative genes and signaling pathways.

## Introduction

Colorectal cancer (CRC) is the fourth most common cancer in the UK and 2^nd^ leading cause of cancer-related fatalities in the UK and US. Over 42 000 people are diagnosed with CRC every year in the UK^1^ and 140 000 in the US. Most CRCs develop from polyps, a pre-cancerous outgrowth of tissue from the lining of the colon. Colonoscopy has long been the reference standard for investigation to examine the entire rectum and colon for precancerous polyps, tumors, or other problems (1,2). A tissue sample (biopsy) taken during colonoscopy helps make a definitive diagnosis of colonic abnormalities. Microscopic examination of colorectal biopsies is a clinical standard to obtain diagnostic information and the key characteristics of CRC tissue for deciding a future management plan, as it may reveal normal findings or a variety of indications of malignancy (1).

The increasing number of patients due to cancer screening programs for early detection adds to pathologist workloads. Early stage disease is more challenging and difficult to detect often increasing the numbers of biopsies as further diagnostic information can be required to provide the best standard of care. Pathology laboratories have seen a doubling of large bowel biopsy slide volumes in the last decade^2^ as opposed to only 10% increase in the number of cases. Digitization of cellular pathology laboratories with increasing ubiquity of digital slide scanners and an accelerated transition to digital pathology during the COVID-19 pandemic (3) offer an opportunity for automated pre-screening of large bowel cancer.

In the clinical workflow, a fundamental diagnostic problem in large bowel cancer screening is the separation of neoplastic from normal biopsies. Normal biopsies are more frequent than neoplastic and form a sizeable portion of the clinical workload. Therefore, it would be clinically beneficial to automate the separation of normal and neoplastic biopsies (4–7). Artificial intelligence (AI) based diagnostic tools for histopathology pre-screening deployed in the digital pathology workflow is an unmet need of clinical research. We conjecture that developing high sensitivity AI models will lead to a reliable and low-risk inclusion of AI-based screening tools in the clinical workflow. An AI-based pre-screening tool will reduce workload and improve impartiality and efficiency. The tool can then be reused and optimized as per the clinical objectives efficiently with incremental, machine aided learning. AI based pre-screening carries the promise of early detection of CRC, which is curable if diagnosed early (1). However, to the best of our knowledge, there is no existing AI tool addressing the screening or pre-screening of colon and rectal biopsies.

In this study, we present an automated method for colon biopsy pre-screening based on digitized whole slide images of colorectal biopsy sections to distinguish between normal and neoplastic cases. A pre-assessment of all biopsies digitally is likely to prove beneficial if done before pathologist evaluation of biopsies. In particular, our study aims at: 1) filtering slides with normal histology out of daily workload; 2) highlighting regions of high neoplastic scores and; 3) neoplastic scoring of each malignant slide which can be used to separate the neoplastic cases into urgent and non-urgent groups for further detailed microscopic examination. Our AI tool can help the efficient management of the time and effort of medical experts for detailed examination of urgent neoplastic biopsies.

In recent years, an increasing number of weakly supervised deep learning methods for whole-slide image (WSI) classification has been proposed for various histopathology problems (8–13). An attractive feature of weakly supervised methods is their ability to enable automatic classification without the need for detailed pixel or regional annotations process. These methods can be made to work efficiently on thousands of WSIs, often after dividing them into smaller parts as image tiles (or patches), during the model training. Recently, Oliveira *et al*. have identified limitations of existing algorithms and underscored the need for more accurate methodologies for use in clinical practice (14). They highlighted the need for larger datasets and the use of appropriate learning methodology to improve prediction accuracy. In our previous work, we proposed an iterative draw and rank sampling based weakly supervised WSI classification pipeline (IDaRS) for prediction of molecular pathways and genetic mutations in CRC (15). For the CRC pre-screening task in this study, we adapt IDaRS for the task of pre-screening of colon biopsies into normal and cancerous categories, termed as the IDaRS-COBI from hereon. We show its potential in data efficiency and its effectiveness for training of a weakly supervised deep learning model. For the task of pre-screening, we demonstrate its high accuracy by cross-validation in a large internal cohort and in an independent validation on a completely unseen external cohort.

## Materials and Methods

### Study Design

The study was undertaken mainly using a cohort of large bowel biopsies scanned at the University Hospitals Coventry and Warwickshire (UHCW), a large academic teaching hospital in the UK providing pathology services for two hospital sites (UHCW and George Eliot Hospital NHS Trust Nuneaton).

During 2019, the cellular pathology laboratory at UHCW processed a total of 41 771 histopathology requests, including 4 877 (11.7%) colon and rectal (large bowel) biopsies of which, 1 680 (34.4%) were normal^3^. The cellular pathology laboratory achieved the milestone of 100% digital scanning of colorectal cases in 2016 with all gastrointestinal pathologists fully validated for digital pathology reporting with the exception of bowel cancer screening cases, which are required to be reported on using glass slides. The sites of biopsies or resection for both UHCW and TCGA cohorts include colon NOS (not otherwise specified), ascending colon, hepatic flexure of colon, caecum, rectosigmoid junction, transverse colon, rectum, descending colon, and sigmoid colon. A total of 12 pathologists were involved in the development of the pre-screening tool. The scale of digitization and active involvement of pathologists was crucial for designing a clinical-grade large-scale study for the development of the proposed AI-based pre-screening tool. To understand the potential workflow implications of the digital pre-screening tool, all colorectal (colonoscopic) biopsies were audited over a 5-year period from 2012 to 2017. The data collected included the colon location and the diagnosis.

### COBI – The development and internal validation cohort

The first cohort of colon biopsies, termed as the COBI cohort, comprises cases from UHCW and is used as the development cohort in this study. All slides in the COBI cohort are diagnostic standard Hematoxylin and Eosin (H&E) stained Formalin-Fixed Paraffin-Embedded (FFPE) histology slides scanned at 40× (0.275 microns per pixels or MPP) using the Omnyx VL120 scanner at UHCW. The slide-level diagnosis for all slides (neoplastic or normal) was through review by at least two pathologists, with additional pathologists providing a consensus diagnosis in instances of discrepancy. Table 1 gives details of the COBI cohort and shows the total number of slides and tiles, and number and percentage of slides and tiles per diagnostic category.

**Table 1.** Cohorts used in this study for development and internal/external validation of the AI pre-screening method. Cases with samples in both diagnostic categories are counted twice.

| Cohort | Diagnostic Categories | Cases | Slides | % | Tiles | % |
| --- | --- | --- | --- | --- | --- | --- |
| **Development & Internal Validation**<br>**(COBI)** | **Normal** | 1 274 | 2 775 | 64.66 | 199 215 | 47.47 |
|  | **Neoplastic** | 907 | 1 517 | 35.34 | 220 484 | 52.53 |
| **Total** |  | **2 181** | **4 292** | **100.0** | **419 699** | **100.0** |
| **External validation**<br>**(TCGA)** | **Normal** | 112 | 134 | 18.33 | 20 709 | 5.81 |
|  | **Neoplastic** | 588 | 596 | 81.67 | 335 488 | 94.19 |
| **Total** |  | **700** | **731** | **100.0** | **356 197** | **100.0** |

### TCGA – Multicentric external validation cohort

The Cancer Genome Atlas (or TCGA) COAD and READ cohorts are used as an external validation cohort in this study. It is a multicentric cohort with slides from 38 different centers in the United States with resections taken during the period 1998–2013. TCGA slides are scanned at 40× (≈ 0.25 micron-per-pixel or MPP, *n*=387) and at 20× (MPP ≈ 0.5, *n*=345). In this cohort, slides with malignant labels (n=597) are all FFPE whereas normal slides (n=135) are frozen sections. In this dataset, in addition to the demographics, genomic characterization information as gene-level copy number variations, mutations and cellular fractions is available for malignant cases.

### Modeling the colon biopsy pre-screening tool

We modeled colorectal biopsy pre-screening as a binary classification machine learning problem where each WSI is classified into one of two categories: normal and neoplastic. Table 2 lists the various types of early signs and malignancies present in colorectal biopsies which characterizes neoplastic category in our development and validation cohorts. Our proposed neoplastic category models early signs and cancerous abnormalities together. It enables the detection of a wide range of malignant conditions and any potential cancer to be monitored and managed from an early stage. Such a sensitive disease pre-screening tool will enhance clinical practice for preventive and proactive disease management. TCGA cohort is used to validate the model’s generalization on an extended range of unseen malignant samples from various centers.

**Table 2.**
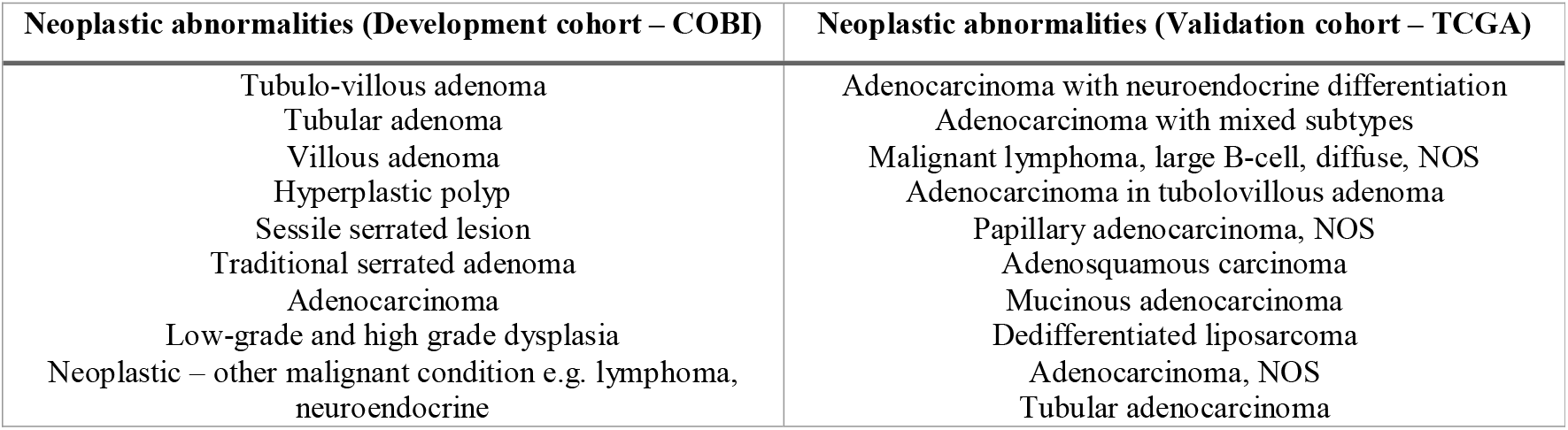
Examples of neoplastic colorectal biopsies in the internal (development) and external (validation) cohorts used in this study.

Figure 1 shows the development and validation methodology of COBI-IDaRS. A deep convolutional neural network (CNN) is trained for binary classification on image tiles from the training slides of the COBI cohort. The prediction scores of image tiles are then aggregated into a single prediction score for the entire slide. The internal validation of the trained model is performed using WSIs from the COBI cohort in a cross-validation setting and external validation using WSIs from TCGA cohort.

**Figure 1.**
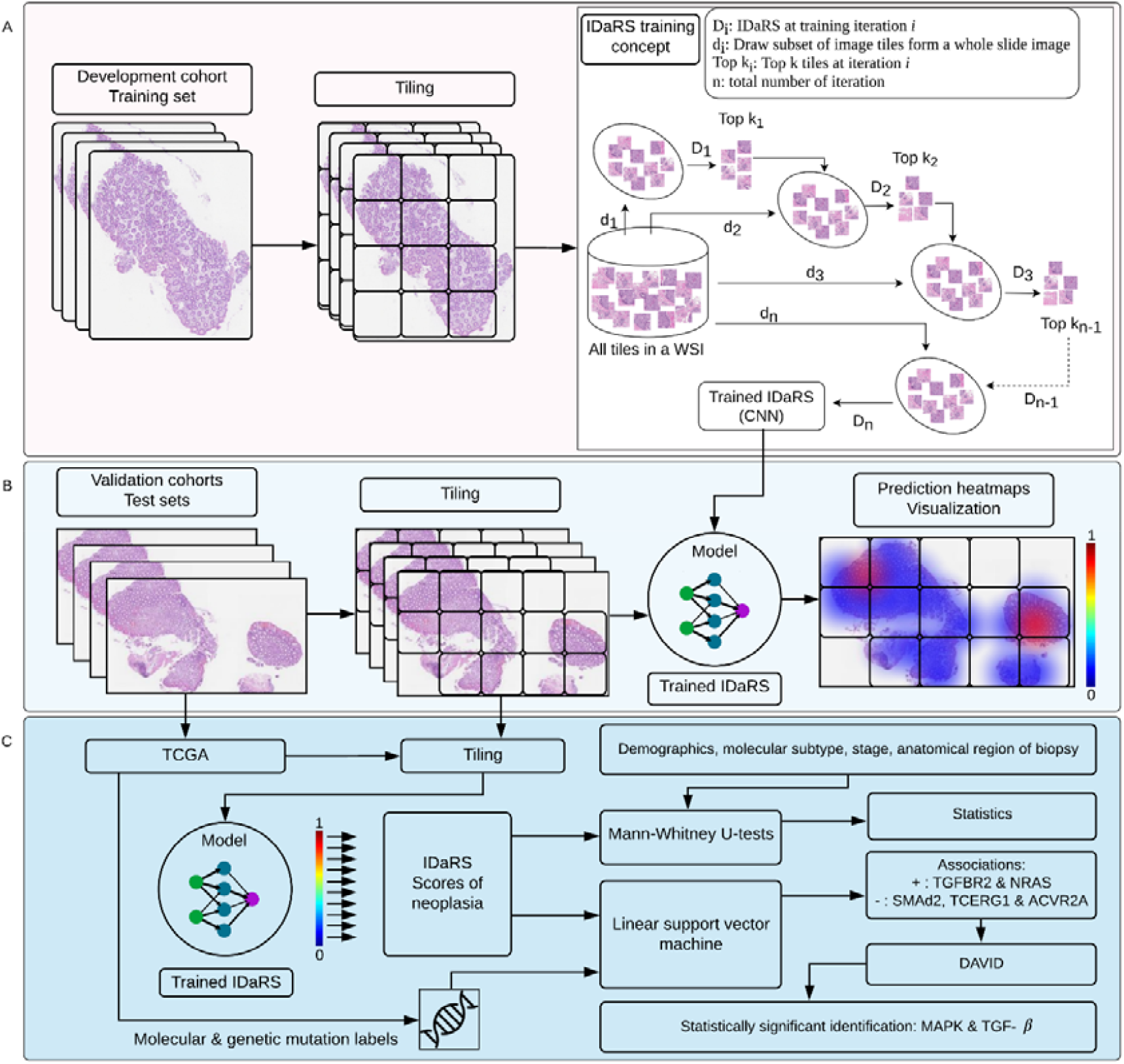
A schematic diagram of the proposed weakly supervised deep learning based tool for pre-screening of colon biopsies: (A) Tissue segmentation and tiling is performed to obtain tiles containing tissue from the slides of development cohort. These tiles serve as input to the iterative draw and rank sampling (an adaptation of ResNet34 model (16)), which was trained on input tiles to learn discriminative features of diagnostic categories. The inset shows a conceptual diagram of the IDaRS training strategy for efficient classification of WSIs. The deep learning model was trained with iterative draw (di) of the fixed percentage of tiles from each WSI while using k of top-ranked tiles of the same slide drawn in the previous iteration; (B) At the time of inference (i.e., validation or testing), the trained IDaRS model gives a prediction score to each tile in the WSI, which are then used to obtain an aggregated slide-level score for each WSI; (C) Validation of IDaRS predicted scores on TCGA slides is achieved via an investigation of their relationship with corresponding genomics and clinical parameters.

### Data preprocessing

Tissue regions are segmented from each WSI to extract tiles corresponding to the tissue areas only. We extracted square tiles of 256×256 pixels from a downscaled version of WSI at a 5× objective magnification (≈2.2 MPP) from the segmented tissue regions. A tile is retained for subsequent processing if the tissue area covers 75% or more of it. WSIs with fewer than four tiles are excluded.

### Weakly supervised deep learning using IDaRS

For training a WSI classifier in a weakly supervised manner, we adapted our published method IDaRS (15), which works on the principle that all image tiles from the tissue regions in a WSI are not equally predictive of the WSI label. Therefore, instead of using all tiles, we choose two subsets of image tiles from each slide for the training. For each training iteration, one subset contains randomly picked proportion (*r*) of tiles and the other contains only the top-ranked proportion (*k*) of the tiles. The parameter settings for IDaRS in this study include *r=10%* and *k=1%*, maximum of T=30 training iterations and batch size of 1024 tiles. We fine-tuned the backbone ResNet34 (16) network (originally pretrained on ImageNet) on the COBI cohort using cross-entropy loss, the Adam optimizer with initial learning rate and weight decay of 10^−4^. During the training iterations, we iteratively use COBI-IDaRS to produce prediction scores for each tile and select the top-ranking *k* tiles of each slide, i.e. those with high likelihood of being neoplastic.

We used PyTorch for the deep learning implementation. A set of data augmentations, including random rotation with maximum angles at 0, 90, 180, and 270 degrees, random horizontal and vertical flip transformations, color jitter with brightness, contrast, saturation of 0.3 and hue of 0.05 were applied on-the-fly on all training tiles (of size 224×224 pixels, ≈2.514 MPP). All experiments were conducted on an Nvidia DGX-2 Deep Learning System with 16× 32GB Tesla V100 Volta GPUs in a shared environment. The deep learning model was built on 2 parallel GPUs with 10 worker threads, with each GPU having a dedicated RAM of 32 GB.

### Experimental setup

We performed three-fold *internal* cross validation with case-controlled stratification for performance evaluation of the proposed pre-screening tool using the COBI cohort. In this evaluation protocol, the dataset is divided into three subsets based on the case identifiers to ensure that all WSIs of a case are in the same fold. For each fold of the cross-validation, two subsets were used for training (with WSIs in the training set randomly split into training and validation sets) while the third served as an unseen *internal* test set. The best performing model, in terms of area under the convex hull of the receiver operating characteristic (AUROC) for the validation set, was chosen as the best performing trained model to obtain predictions on the held-out test set in a blinded manner.

For performance evaluation, we use the following metrics: AUROC, the average precision of precision-recall curve (AUPRC), sensitivity and specificity at three selected thresholds. All evaluation metrics are averaged for multiple runs and multiple folds of each experiment. We report the mean and standard deviation of AUROC, as well as those for AUPRC. Moreover, we investigated the average specificity values at sensitivities of 0.95, 0.97, and 0.99, as the benchmarks to evaluate the effectiveness and robustness of the AI tool to serve in a real clinical setting. In the Supplementary materials, (Table S2, p.2) shows details of the data folds used for 3-fold cross validation experiment on the COBI development cohort. It gives number of cases, slides, and tiles for each cohort, class, set and fold.

### Demographic and mutation characterization analysis

Malignant samples in the TCGA cohort have associated information about demographics (gender and race), molecular subtype, stage, anatomical region of biopsy, mutation genotypes as well as immune expression signatures (17). We performed statistical analysis to identify any systematic differences between model-generated probability values or prediction scores for a test case being classified as cancerous based on these factors. For this purpose, we employed the non-parametric Mann-Whitney U-tests between a factor of interest (such as gender or stage, etc.) and the prediction score generated by the proposed model.

We also estimated the association between occurrence of different colorectal gene mutations and the prediction score for malignant cases obtained from the model. Any such association is expected to indicate possible preferential scoring by the model for a given set of gene mutations, as well as signaling pathways implicated in colorectal cancer. We analyzed the bootstrap average of weights of a linear support vector machine classifier trained to predict whether the prediction score of a given case is above or below the median value of prediction scores across all cases using gene mutation information for that case alone. The resulting estimate can be regarded as the relative association of mutation in a given gene with the model prediction score. We also performed signaling pathway enrichment analysis using DAVID (18,19) based on genes whose mutations receive a large magnitude of average weights across 100 bootstrap runs.

## Results

The IDaRS-COBI gives two probabilities to each tile at the test time. These probabilities correspond to the likelihood of a tile belonging to normal and neoplastic histology in the binary classification setting. Scores of various numbers of tiles in a WSI are considered for aggregation with multiple schemes into a single WSI score, which represent likelihood scores (probabilities) of WSIs being neoplastic. The performance metrics AUROC, AUPRC measures, sensitivities, and specificities are obtained from the WSI scores. To generate the slide-level label, IDaRS-COBI employs the “Average Top Half” aggregation scheme, which calculates an average of prediction scores of tiles with score higher than the median score of all tiles in the slide.

Mean and standard deviation values of all performance metrics are reported for an unseen test set in a 3-fold *internal* cross-validation on the COBI cohort (n=4 292) and *external* validation on the TCGA cohort (n=731), as shown in Table 3. For comparative analysis of the IDaRS-COBI results, we also report same performance measures of a recent state-of-the-art data-efficient method named CLAM (8), for the two cohorts with the same training and testing splits. It can be seen from Table 3 that IDaRS-COBI produced higher accuracy values on all reported measures. We report better results of CLAM obtained using all training data, parallel attention branches (8), and keeping the subtype parameter active. For IDaRS-COBI, in the Supplementary materials (Table S1, p.1), we also report results using different aggregation schemes of maximum probability, weighted average probability, and simple average of all probabilities, average of top four and top ten probabilities, from each slide.

**Table 3.** Quantitative performance evaluation of the proposed pre-screening tool (IDaRS-COBI) as compared to another recently proposed method (CLAM) for both internal (COBI) and external (TCGA) cohorts.

| Methodology | Average AUROC | Average AUPRC | Sensitivity 0.95 | Sensitivity 0.97 | Sensitivity 0.99 |
| --- | --- | --- | --- | --- | --- |
|  |  |  | Specificity |  |  |
| Internal 3-fold cross-validation on the COBI cohort |  |  |  |  |  |
| CLAM (8) | 0.9836 (0.0011) | 0.9804 (0.0017) | 0.9478 (0.0073) | 0.8864 (0.0036) | 0.5183 (0.1513) |
| IDaRS-COBI | 0.9895 (0.0021) | 0.9851 (0.0017) | 0.9543 (0.0102) | 0.9173 (0.0201) | 0.7986 (0.0718) |
| External evaluation (TCGA cohort) of all models obtained in 3-fold cross validation (COBI cohort) |  |  |  |  |  |
| CLAM (8) | 0.8819 (0.0444) | 0.9662 (0.013) | 0.5 (0.202) | 0.4154 (0.2123) | 0.2761 (0.1748) |
| IDaRS-COBI | 0.9746 (0.0159) | 0.9945 (0.0034) | 0.8524 (0.1662) | 0.738 (0.1947) | 0.4768 (0.1886) |
| Trained on entire COBI cohort and tested on TCGA cohort |  |  |  |  |  |
| IDaRS-COBI (Validation/TCGA) | 0.9708 | 0.9932 | 0.8806 | 0.7687 | 0.3955 |

Figure 2a shows the shaded ROC curve while Figure 2b the sensitivity-vs-specificity curve at sensitivity values between 0.8 and 1.0. It can be observed that at the sensitivity value of 0.99, IDaRS-COBI gives a specificity of 0.80 for the COBI cohort. Higher specificity can be achieved if the sensitivity threshold is relaxed to 0.97 (specificity = 0.92), or 0.95 (specificity = 0.954). The ROC and sensitivity-vs-specificity plots (for sensitivity > 0.8) for external validation on TCGA cohort are shown in Figure 2c and Figure 2d. It is perhaps worth noting that the normal tissue sections in the TCGA cohort were frozen and from the adjacent normal tissue and, therefore, it is possible that the model picks more than just the neoplastic histology. Being that in mind, for TCGA cohort, we extended our analysis to further explore the neoplastic regions as identified by the model within the neoplastic H&E slides. We investigated digital scores predicted by IDaRS-COBI in neoplastic regions and the association of our digital scores with relevant clinical and molecular variables of neoplastic samples in the TCGA cohort, as reported below (see section titled Results of Demographic and Mutation Characterization Analysis).

**Figure 2.**
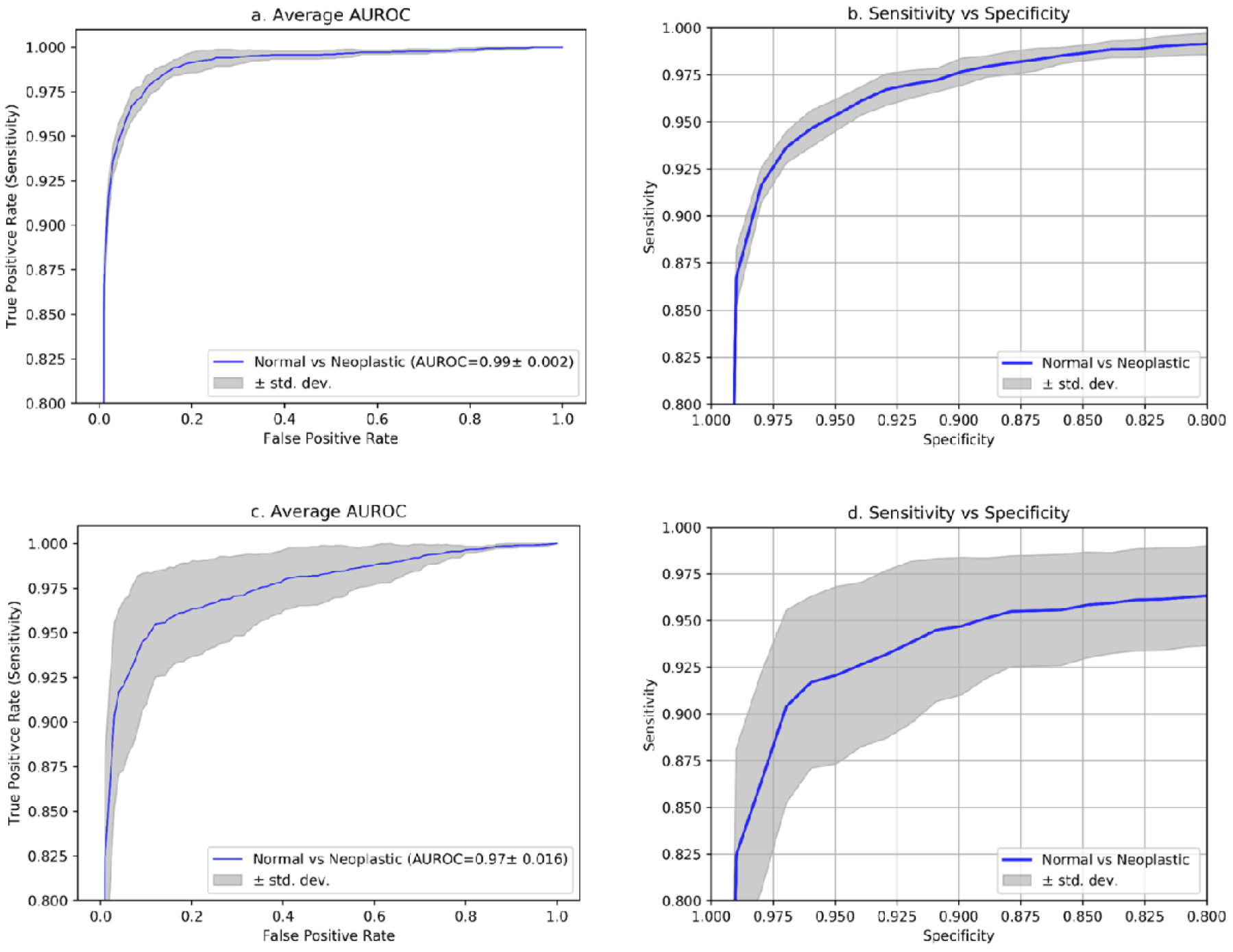
ROC plots and sensitivity vs specificity plots of the proposed IDaRS-COBI method on a 3-fold internal cross-validation on the COBI cohort (a, b) and external validation on the TCGA cohort (c, d).

Figure 3 shows input image tiles (a – d) of accurately predicted tiles for neoplastic cases and the corresponding saliency maps (e – h) overlaid on the input images. Saliency maps here highlight all pixels contributing to the prediction of neoplastic label for the given image tiles. It is worth noting that image tiles corresponding to different malignancy subtypes (hyperplastic polyp, poorly differentiated adenocarcinoma, dysplasia/adenocarcinoma, high grade invasive carcinoma, as shown in Figure 3a-d) are predicted correctly at the tile level by the model and verified by a pathologist (YWT) using saliency maps.

**Figure 3.**
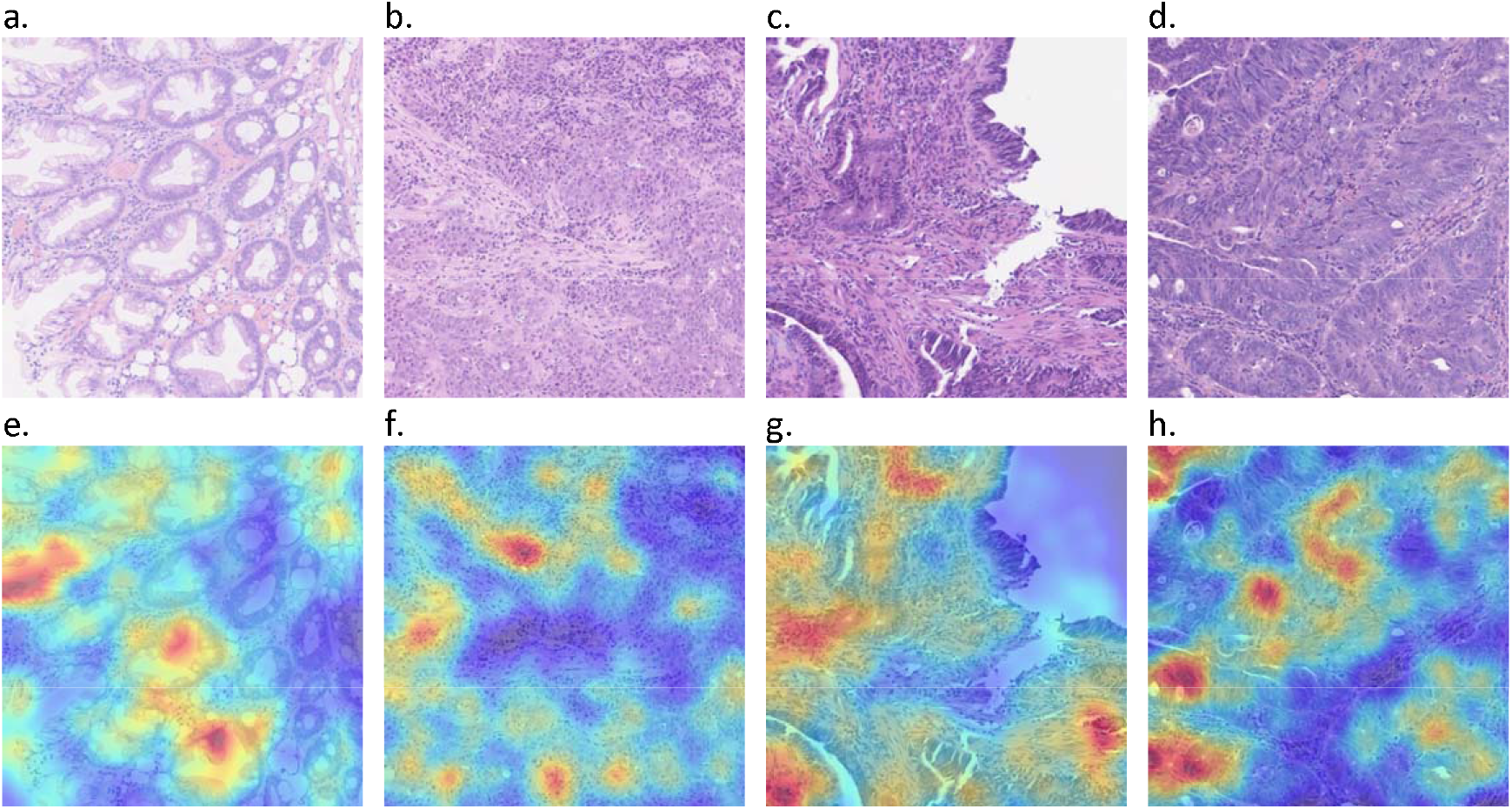
Example image tiles of accurately predicted biopsies of a Hyperplastic polyp (a), adenocarcinoma (b), dysplasia/adenocarcinoma (c) and high grade dysplasia (d) from neoplastic colonic (a, b) and rectal (c, d) biopsies and their corresponding saliency maps (e-h) showing regions which contribute towards the accurate predictions. Gaussian smoothing has been applied on the saliency maps for improved visualization.

Sample image tiles of the normal slides incorrectly predicted with high probability of being neoplastic are shown in Figure 4. The model correctly identified image tiles of hyperplastic polyps (a-c) and tubular adenoma (d, e) and incorrectly identified normal small bowel (l) as neoplastic. The remainder of the images (f-k) show variation of normal, reactive changes, crypt distortion and artifact, which are wrongly attributed high probability of being neoplastic by the model. Sample normal image tiles correctly identified from the normal and neoplastic slides are also shown in Figure S1 (Supplementary p.2). Since we applied a weakly supervised classification for training IDaRS-COBI, all tiles from a slide were given the same slide label during training which will have added the noise in to the trained predictor.

**Figure 4.**
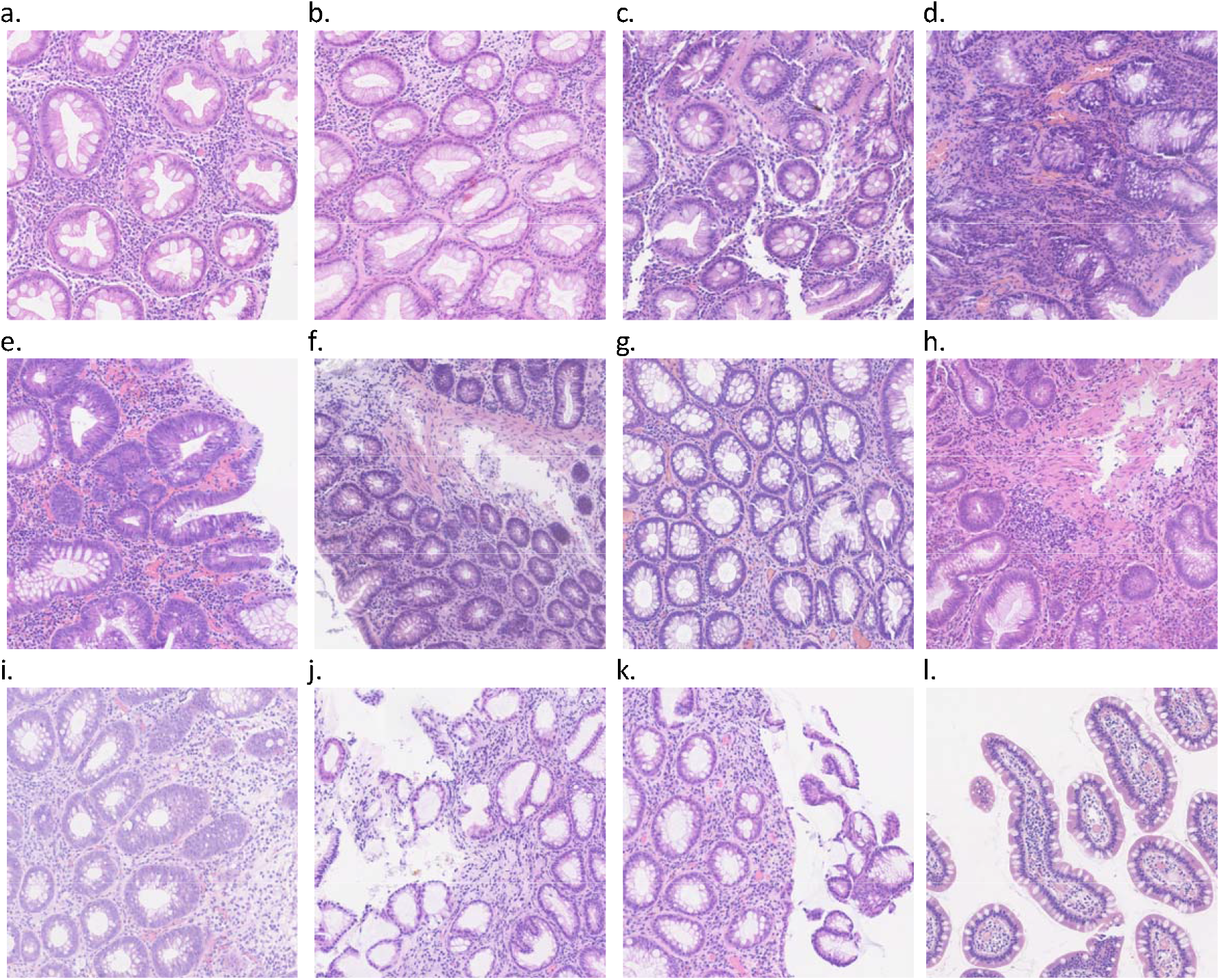
Example image tiles predicted as neoplastic within normal slides of the COBI test set: hyperplastic polyps (a-c), tubular adenoma (d, e), variation of normal, reactive changes, and crypt distortion (f-k), and normal small bowel (l).

Figure 5 shows the prediction heatmaps overlaid on the input images with a corresponding sample image tile for a normal (a) and a neoplastic slide (b), respectively. As can be seen in Figure 5a, all the tiles are predicted as normal (shown by the blue overlay), confirming normal histology. Similarly, as can be seen in Figure 5b, most tiles are predicted as neoplastic (shown by the red overlay) confirming neoplastic histology. More examples of neoplastic slides with their corresponding overlay heatmaps can be found in supplementary figures Figure S2 and Figure S3 (Supplementary, p.2). Prediction of normal tiles in most of normal cases matches with the pathologist views. A few cases of sample normal histology tiles predicted as neoplastic in the neoplastic slides are shown in Figures S2 and S3. As our method did not use any regional or cellular annotations for training the prediction model, this can be expected. However, we would like to argue that such minor errors have minimal impact on the clinical workflow as long as the final WSI-level score points to a correct diagnostic category. Figure S4a (Supplementary, p.3) shows an interesting slide categorized as normal in the given ground truth labels whereas the proposed algorithm labeled it as neoplastic. The algorithm ‘s prediction was corroborated by our team of pathologists when they reviewed the slide and its corresponding heatmap (as in Figure S4b) and images of top-ranked tiles. The algorithm was also able to pick up other slides that were originally labeled as normal but were predicted as neoplastic by the algorithm, also further attested by a consensus of our pathologists.

**Figure 5.**
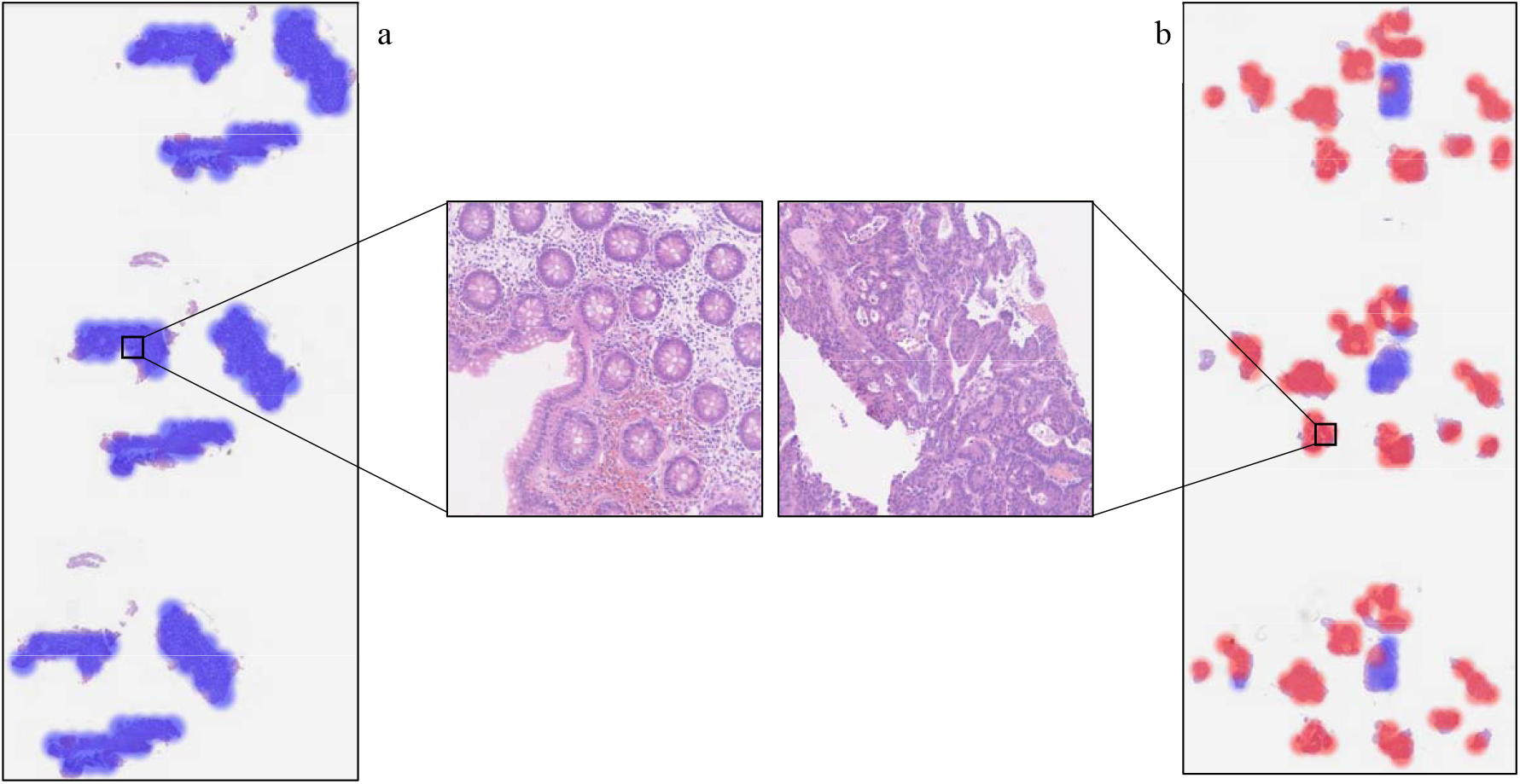
Heatmap visualizations of COBI slides. a. Heatmap overlay of normal slide and a sample normal tile. b. neoplastic slide with a sample neoplastic tile. Overlay color legend: blue (normal) and red (neoplastic).

### Results of Demographic and Mutation Characterization Analysis

Our analysis revealed no statistically significant differences across prediction scores from the model for malignant cases based on their gender (Male/Female), stage (I, II, III or IV) or anatomical site of the biopsy (Descending, Ascending or Transverse Colon or Rectum) with Mann-Whitney U-test p-values > 0.05. Also, no statistically significant differences between prediction scores for malignant cases were observed based on specific molecular subtypes (Chromosomal Instability (*p* = 0.59), Genomic Stability (*p* = 0.02) or Hypermutated Single Nucleotide Variants (*p* = 0.43)). However, the average prediction scores for Microsatellite Instability High cases (MSI-H) was found to be slightly lower in comparison to other groups (Mann Whitney U-test p-value of 0.0002). Similarly, a statistically significant difference between prediction scores for Caucasian cancer cases (n = 243) in the TCGA set in comparison to other demographics was observed (*p* < 0.0005). However, the effect sizes for these differences are practically insignificant indicating the future need for a larger validation cohort to confirm any such potential biases.

Our analysis of association between prediction scores and gene mutations revealed that mutations in the TGFBR2 and NRAS genes were positively associated with model prediction score whereas those in SMAD2, TCERG1 and ACVR2A genes were negatively associated with the prediction score (see Supplementary Figure S5, p.3). Gene set enrichment analysis (GSEA) over these genes through DAVID resulted in the statistically significant identification (*p* < 0.05) of MAPK and TGF-β signaling pathways as being associated with increased and decreased prediction scores from the model, respectively (see supplementary Figures S6-S8, p.3 and p.4).

Figure S5 (Supplementary p.3) shows the average weights of different genes from the SVM regression analysis against a model prediction score plotted against the probability of mutation occurrence in cancerous cases in the TCGA cohort, pointing to the role of important genes (red stars) whose prediction scores are associated with the model prediction scores obtained from DAVID Gene Set Enrichment Analysis.

## Discussion

In this study, we have proposed a weakly supervised AI histology image pre-screening tool for CRC based on a variant of the IDaRS algorithm (15). The method presented in this study is aimed at assisting large bowel biopsy screening in clinical practice with a prior digital pre-screening. Our results demonstrate that AI based pre-screening offers the potential for improved efficiency as well as improved reliability.

The proposed IDaRS-COBI tool yielded sensitivity values of 0.99, 0.97, and 0.95 for normal vs neoplastic slides, with corresponding specificities of 0.80, 0.92, and 0.95, respectively. Picking a highly sensitive operating point (with sensitivity of 99%) can help greatly reduce the pathologists’ workload by automated reporting of normal cases allowing their time to be spent examining more complex cases. The proposed AI tool has also been shown to be capable of identifying cancerous regions from all tumor slides of the multicentric TCGA cohort used for the external validation.

To deploy in practical histopathological diagnostic workflow system for screening colon biopsies, a recent study (20) modeled the classification of epithelial tumors (adenocarcinoma and adenoma) with a large development cohort (n=4 036) and small validation cohorts (n=500, and TCGA-COAD: n=547). A more recent study (21) used gland segmentation and classification to model categorization of ‘low risk’ (benign, inflammation) and ‘high risk’ (dysplasia, malignancy) slides with a small cohort (n=294). However, these models do not cater for early diagnosis and lack high sensitivity. We designed a sensitive AI model that can differentiate between neoplastic and normal large bowel biopsies while being able to pick up hyperplastic polyps and dysplastic lesions, thus paving the way for a reliable and more efficient pre-screening step in the routine clinical workflow with the promise of reduced pathologist workload.

The performance of our AI tool outperforms recently proposed supervised and semi-supervised AI tools (4,22) for CRC diagnosis considering multiple aspects. Wang *et al*. in (4) proposed a fully supervised AI tool for cancer vs. normal prediction while Yu *et al*. (22) proposed a semi-supervised AI tool which matches to the performance of supervised AI. In both, semi and fully supervised classification authors have required both the slide-level and laborious tile-level annotations to achieve a sensitivity of 0.982, showing a slight improvement over an average performance of six pathologists (sensitivity = 0.975) on the same test set (4). However, it is not desirable for an AI pre-screening tool to yield high specificity at the cost of sensitivity. Our proposed weakly supervised algorithm improves sensitivity up to 0.99 at the cost of slightly reduced specificity (0.80). The proposed algorithm does not require any regional or cell level annotations, thereby saving the time and cost of laborious manual annotation. We have shown that the algorithm was able to identify a few errors in the slide-level ground truth labels, which may be linked to human errors. The algorithm can also pick sparse features of neoplasia and early signs of abnormalities, which may be overlooked under the microscope given the vast amount of tissue content to be analyzed manually.

To mitigate the effect of tile-level mispredictions, we considered different aggregation schemes to combine tile-level probability scores into slide-level probability scores and effectively filter out noise in tile-level predictions. As an additional training strategy, we also used a third subset of the top normal tiles from all slides during IDaRS-COBI training. This subset contained 0.5% of top normal tiles from normal and neoplastic slides added in the training set by pseudo-labelling normal tiles of neoplastic slides as normal. We noticed an improvement in cross-validation performance of IDaRS-COBI on the development cohort (AUROC=0.9902±0.0013, AUPRC=0.9854±0.0018, Specificity=0.8282±0.0297 at sensitivity of 0.99). We did not use explainable cellular features or richly annotated image tiles to build the deep learning model. We relied on automatic feature extraction of the image tile-based weakly supervised deep learning, because of its efficient processing, automatic feature engineering and not necessitating laborious detailed annotations. However, we investigated the automatically learned features by saliency map visualizations which led to identifying a few false positives at tile levels as well as errors in the ground truth labels. Some of the false positives were found to be insignificant or irrelevant features and mimickers amplified in the neoplastic tissues. The false positives at tile levels have been mitigated with a selection of better aggregation schemes. Moreover, assigning a single ground-truth label to all tiles from multiple slides belonging to the same case may be interpreted as *overfitting* in classical machine learning theory. Despite the tile-level false positives, the quantitative measures have shown excellent predictive accuracy, robust performance and generalization on both cohorts. This may be associated with interpolation and generalization capabilities of overparameterized deep learning systems leading to *benign overfitting*, as demonstrated by Bartlett *et al*. in their latest findings (23).

Additionally, based on the genetic mutations and gene enrichment analysis on external validation cohort, it can be hypothesized that the proposed model generates larger prediction scores for cases with increased MAPK signaling in comparison to those with TGF-β pathway activity. This would be in line with the known roles of these pathways in colorectal cancer (24). A statistically significant negative Pearson correlation of −0.18 (*p* < 0.00001) between prediction scores and the TGF-β response values obtained from (17) also lends support to these findings. The statistically significant Pearson correlation of 0.16 (*p* < 0.005) between prediction scores and the estimated values of absolute tumor purity (percentage of cancer cells in a solid tumor sample) from (17) also indicates that the model is possibly more sensitive towards detection of proliferation-centered mechanisms in colorectal cancer cases.

A weakly supervised model with sensitivity greater than 99% and higher level of specificity is achievable with a large-scale, international, multi-cohort validation while ensuring the slide-level diagnostic labels are correct. Such an outcome is highly likely to assist with screening of colorectal biopsies for multiple abnormalities in a clinical setting. As mentioned above, a digital pre-screening tool such as IDaRS-COBI can result in time and cost efficiency, reliability and higher accuracy of routine diagnostic screening by filtering out normal colorectal biopsies from the daily workload of the pathologist and highlighting the neoplastic regions on the slide. Furthermore, high-scoring true positive cases can also be pre-screened digitally and prioritized by an expert ‘s examination to further improve the management of clinical workload. Finally, we argue that digital biopsy pre-screening for colon and rectal screening is the need of the day, which connects modern digital technologies and human expertise together consequently enabling significant decrease in cancer fatalities and under- or over-treatment as a result of timely diagnosis of cancer.

Limitations of this study include a class imbalance in the dataset, which is often the case in medical datasets and which, it may be argued, overrate AUROC values. To mitigate the effect of class imbalance, we have also reported AUPRC measure as well. We also expect that after careful revision of the ground truth labels and introducing an additional supervisory signal for model training, both sensitivity and specificity of the model should be further improved. As noted above, the external validation on multicentric TCGA cohort has a caveat that all normal slides from that cohort are of frozen sections. However, being able to identify neoplastic tiles in all H&E neoplastic slides of TCGA cohort is an encouraging outcome of the external validation in addition to demonstrating the generalization ability in an independent cross-validation setting. In future, these results merit validation on a large and diverse multicentric cohort, paving the way for improving the model performance and evidencing generalization before its eventual clinical deployment.

## Data Availability

The annotations and the COBI cohort will be made available upon completion of the PathLAKE project. All images and the clinical, demographic, and mutation status information for the TCGA COAD and READ cohort used in this study are publicly available at https://portal.gdc.cancer.gov/ and cBioPortal (https://www.cbioportal.org/).

## Acknowledgments

Manuel Salto-Tellez and Jacqueline A James are Principal Investigators in PathLAKE at Queens ‘s University Belfast, and Clare Verrill is a Principal Investigator in PathLAKE at the University of Oxford – all were involved in generating the PathLAKE Programme, including funding.

This paper is supported by the PathLAKE Centre of Excellence for digital pathology and artificial intelligence, which is funded from the Data to Early Diagnosis and Precision Medicine strand of the HM Government ‘s Industrial Strategy Challenge Fund, managed and delivered by Innovate UK on behalf of UK Research and Innovation (UKRI). The views expressed are those of the authors and not necessarily those of the PathLAKE Consortium members, the NHS, Innovate UK or UKRI.

The results we report are in part based on data generated by The Cancer Genome Atlas Research Network. The authors would like to thank Johnathan Breddy, Young Park, Giorgos Hadjigeorghiou, John Euinton, Chien-Hui Liao, Thomas Leech and Valentine Stoykov for their assistance with administrative matters and IT support for software and systems utilized heavily in this project.

## Ethics Approval and Consent to Participate

This study was conducted under Health Research Authority National Research Ethics approval 15/NW/0843; IRAS 189095 and the Pathology image data Lake for Analytics, Knowledge and Education (PathLAKE) research ethics committee approval (REC reference 19/SC/0363, IRAS project ID 257932, South Central - Oxford C Research Ethics Committee).

## Authors contributions

MB, DS and NR designed the study with support from all co-authors. MB, FM and NR developed the methods. MB wrote the code and carried out all experiments. YWT, KG, MA, EH, AR, AA, EH, KD, HS, MN, KB and DS provided diagnostic annotations of colonic biopsy slides. YWT, MB, FM, KG, MA, DS and NR performed analysis and interpretation of the results. FM performed demographic and mutation characteristic analysis on the TCGA cohort. SEAR, AB, SG, MJ, NW and WL provided technical and material support. MB, YWT, FM, DS and NR were all involved in drafting of the paper; All authors read and approved the final paper.

## Footnotes

1 https://www.bowelcanceruk.org.uk/about-bowel-cancer/bowel-cancer/

2 Based on data from the internal UHCW pathology laboratory requests over the last decade.

3 The data from 2019 shows a doubling of case volume as compared to 2008. The percentages may vary among different hospitals and for different years but nevertheless give an indication of the reporting workload.

